# Depression and loneliness during COVID-19 restrictions in the United States, and their associations with frequency of social and sexual connections

**DOI:** 10.1101/2020.05.18.20101840

**Authors:** Molly Rosenberg, Maya Luetke, Devon Hensel, Sina Kianersi, Tsung-chieh Fu, Debby Herbenick

**Author notes:** Corresponding author: Molly Rosenberg 1025 E. 7^th^ Street, Office C032 Bloomington, IN 47405.

## Abstract

**Purpose:** To estimate the prevalence of depression and loneliness during the US COVID-19 response, and examine their associations with frequency of social and sexual connections.

**Methods:** We conducted an online cross-sectional survey of a nationally representative sample of American adults (n=1010), aged 18-94, running from April 10-20, 2020. We assessed depressive symptoms (CES-D-10 scale), loneliness (UCLA 3-Item Loneliness scale), and frequency of in-person and remote social connections (4 items, e.g. hugging family member, video chats) and sexual connections (4 items, e.g. partnered sexual activity, dating app use).

**Results:** One-third of participants (32%) reported depressive symptoms, and loneliness was high [mean (SD): 4.4 (1.7)]. Those with depressive symptoms were more likely to be women, age 20-29, unmarried, and low-income. Frequent in-person connections were associated with lower depression and loneliness; frequent remote connections were not.

**Conclusions:** Depression and loneliness were elevated during the early US COVID-19 response. Those who maintained frequent in-person, but not remote, social and sexual connections had better mental health outcomes. While COVID-19 social restrictions remain necessary, it will be critical to expand mental health services to serve those most at-risk and identify effective ways of maintaining social and sexual connections from a distance.

## INTRODUCTION

The spread of coronavirus disease 2019 (COVID-19) has led to unprecedented changes in the daily lives of Americans. Measures to mitigate the spread of COVID-19, including social distancing and sheltering in place, limit people’s ability to maintain social, relational, and sexual connections and, accordingly, may have dramatic consequences people’s psychological well-being.[1, 2] Social isolation, which is essentially mandated to reduce transmission of coronavirus, has been previously linked to loneliness and negative mental health outcomes in numerous populations.[3, 4] Further, recent research has shown that the spread of COVID-19 and the related public health response have had negative effects on people’s emotional well-being,[5–8] with more than one-third of American adults reporting serious changes in their mental health due to the COVID-19 spread and response.[9, 10]

Under normal conditions, depression is common worldwide and has a significant health burden, affecting some 264 million people each year, according to the World Health Organization.[11] Loneliness is also prevalent and strongly correlated with adverse mental and physical health outcomes, including depression.[12] In the United States, depression has increased among adults over the last decade,[13] largely among young adults and women.[14] Recent estimates from the National Institute of Mental Health indicate that 17.3 million American adults (7.1%) experienced a major depressive episode in 2017.[15] Loneliness tends to follow similar demographic trends as depression, with elevated levels observed in women and young adults, but also among low-income and unmarried people. [12, 16]

While previous studies have evaluated the psychological consequences of other respiratory disease epidemics,[17–20] few studies have been conducted to assess these outcomes related to the current pandemic of coronavirus (COVID-19).[5–7] To our knowledge, no COVID-19-era study has yet estimated the population-representative prevalence of depression and loneliness in American adults. Further, there is limited understanding of how social and sexual connections are associated with mental well-being amidst a global pandemic and the associated containment measures such as quarantine, social distancing, and self-isolation.

This study fills these knowledge gaps by estimating the population-representative prevalence of depression and loneliness in American adults during the early stages of the COVID-19 spread and restrictions, and by examining the frequency of social and sexual connections as potential predictors of these mental health outcomes.

## METHODS

### Study setting, participants, and data collection

We conducted a nationally representative on-line survey of American adults (age 18+ years) from April 10–20, 2020 – the 2020 National Survey of Sexual and Reproductive Health During COVID-19. Participants were recruited into the study using the Ipsos KnowledgePanel, a nationally representative, probability-based sample established using address-based sampling via the U.S. Postal Service’s Delivery Sequence File, of which other surveys have used to underpin high-quality and generalizable results.[21–23] Using an equal probability selection method, members of the panel were sampled and invited to participate in our study. Sampled participants were emailed an invitation and link to an on-line survey querying information on mental health, relationships, sexual behaviors. Ipsos maintains an incentive program for participation in individual surveys, including sweepstakes/raffles for prizes and cash rewards. Ethical approval for the study protocol was provided by the Indiana University Human Subjects Office (#2004194314).

### Key variables

We assessed two key outcomes: depression and loneliness. We measured depression with the 10-item Center for Epidemiologic Studies Depression Scale.[24] As in other studies,[25, 26] we used a scale cutpoint of greater than or equal to 10 to identify those likely experiencing significant depressive symptoms. We measured loneliness with the UCLA 3-item Loneliness scale.[27] We calculated the continuous scale score (possible range 3–9), and used a score of 4 or greater to define a dichotomous loneliness category for comparison with prior studies.[28, 29] We used a more conservative loneliness definition (score of 6 or greater) for categorization for subsequent analyses. This categorization decision was made to address concerns about the likely lower discriminatory value of one of the scale items (‘How often do you feel isolated from others?’).

We assessed the frequency of eight social and sexual connection exposures with reference to the time frame of the last month, each with response categories: not at all, once or a few times, 1–3 times a week, and almost every day. We grouped the social connections into in-person (hug/kiss a family member, visit a non-household friend or family member) and remote (video chat with friend/family, send/receive a letter in the mail). Similarly, we grouped the sexual connections into in-person (hug/kiss a partner, partnered sexual activities: mutual masturbation, oral sex, or intercourse) and remote (sex over phone, video chat or texting; use of dating or hookup app). Few endorsed these final two remote sexual exposures at high frequency, so for those we combined the ‘1–3 times a week’ and ‘almost every day’ responses. We also assessed relationship tension among participants reporting romantic relationship. We measured romantic tension with an item that queried whether participants noted increased tension or arguments with their partner due to COVID-19 spread and restrictions. We categorized responses into ‘yes’ (often, sometimes, or rarely) and ‘no’ (no significant change, or feeling more connected).

We assessed other key demographic variables with data maintained in the Ipsos KnowledgePanel: gender; age, in years, categorized in 10-year increments; race/ethnicity; marital status; region: northeast, south, Midwest, west; household size, and household makeup (living alone, children under 13 years in home). To assess the potential importance of living with a romantic partner, we created a dichotomous *relationship status* variable separating participants who lived with a romantic partner from participants who did not live with a romantic partner, either because their romantic partner lived elsewhere or because they did not have a romantic partner.

### Statistical analysis

To account for any differential nonresponse that may have produced different distributions between our survey sample and the overall Ipsos KnowledgePanel, all analyses were conducted on a sample weighted to the overall to the geo-demographic distribution of the United States based on US Census data. The magnitude of missing data was <5% for all variables, so we conducted complete case analyses. We calculated descriptive estimates of the prevalence of depressive symptoms and loneliness, and used chi-square and t-tests to assess which categorical and continuous demographic variables differed significantly by depression status. We used log-binomial regression to estimate the associations between each of the nine exposures (frequency of social and sexual connections and relationship tension) and each of the two mental health outcomes (depression and loneliness). We estimated these associations in unadjusted models, and in models adjusted for gender, age, marital status, and income. To examine whether the associations differed by relationship status, we added an interaction term between relationship status and each of the nine exposures, dichotomizing the frequency variables at 1–3 times per week (at or above/below). We estimated stratified results and used the magnitude of differences in point estimates and Wald p-values to inform our conclusions around effect measure modification. All analyses were conducted using SAS statistical software, version 9.4.

## RESULTS

Of the 1632 KnowledgePanel members invited to participate in the study, 1010 completed the survey for a 61.9% response rate. The weighted sample was 52% female, 63% non-Hispanic White, 12% non-Hispanic Black, 16% Hispanic, and 9% other race or multiple races. Most (62%) were married, and one-third (33%) had a college education or higher (Table 1). The demographic differences between the weighted and the unweighted sample were minor. The ages of those in the sample ranged from 18 to 94 years and the weighted median age was 48 years (IQR 32–62).

**Table 1.** Characteristics of the unweighted and weighted sample, overall and by depressive
symptoms in 1010 American adults, April 10–20, 2020

| Characteristic | Unweighted<br>Overall<br>N=1010 | Weighted<br>Overall<br>N=1010 | Major<br>depressive<br>symptoms<sup>1</sup><br>N=304 | Below<br>threshold for<br>major<br>depressive<br>symptoms<sup>1</sup><br>N=657 | p-value |
| --- | --- | --- | --- | --- | --- |
|  | N (%) | N (%) | N (%) | N (%) |  |
| **Sex** |  |  |  |  | 0.002 |
| Female | 516 (51.1) | 521 (51.6) | 179 (58.9) | 315 (47.9) |  |
| Male | 494 (48.9) | 489 (48.4) | 125 (41.1) | 342 (52.1) |  |
| **Age group** |  |  |  |  | <0.0001 |
| 18-19 | 15 (1.5) | 27 (2.7) | 10 (3.4) | 14 (2.2) |  |
| 20-29 | 114 (11.3) | 184 (18.2) | 76 (24.9) | 99 (15.1) |  |
| 30-39 | 156 (15.5) | 175 (17.4) | 55 (17.9) | 106 (16.1) |  |
| 40-49 | 138 (13.7) | 142 (14.1) | 45 (14.6) | 92 (14.0) |  |
| 50-59 | 215 (21.3) | 185 (18.3) | 59 (19.4) | 119 (18.1) |  |
| 60-69 | 206 (20.4) | 165 (16.3) | 33 (10.9) | 126 (19.3) |  |
| 70-79 | 129 (12.8) | 103 (10.2) | 23 (7.6) | 76 (11.6) |  |
| 80+ | 37 (3.7) | 28 (2.8) | 3 (1.1) | 24 (3.7) |  |
| **Race/ethnicity** |  |  |  |  | 0.3 |
| White, non-Hispanic | 721 (71.4) | 639 (63.2) | 182 (59.7) | 434 (66.1) |  |
| Black, non-Hispanic | 85 (8.4) | 119 (11.8) | 41 (13.6) | 71 (10.8) |  |
| Other or 2+ races, non-Hispanic | 81 (8.0) | 87 (8.6) | 29 (9.6) | 53 (8.1) |  |
| Hispanic | 123 (12.2) | 165 (16.4) | 52 (17.2) | 99 (15.1) |  |
| **Marital status** |  |  |  |  | <0.0001 |
| Married or living with partner | 668 (66.1) | 622 (61.6) | 152 (50.0) | 444 (67.5) |  |
| Separated/divorced | 129 (12.8) | 124 (12.3) | 40 (13.1) | 76 (11.6) |  |
| Widowed | 44 (4.4) | 40 (3.6) | 14 (4.6) | 23 (3.6) |  |
| Never married | 169 (16.7) | 224 (22.2) | 98 (32.3) | 114 (17.3) |  |
| **Region** |  |  |  |  | 0.8 |
| Northeast | 189 (18.7) | 177 (17.5) | 52 (17.1) | 111 (17.0) |  |
| Midwest | 228 (22.6) | 210 (20.8) | 66 (21.6) | 134 (20.4) |  |
| South | 341 (33.8) | 383 (37.9) | 110 (36.1) | 258 (39.2) |  |
| West | 252 (25.0) | 240 (23.8) | 77 (25.2) | 154 (23.4) |  |
| **Household income** |  |  |  |  | 0.0005 |
| <\$25k | 95 (9.4) | 136 (13.5) | 61 (20.2) | 69 (10.6) | |
| \$25-49k | 174 (17.2) | 184 (18.2) | 54 (17.7) | 120 (18.3) | |
| \$50-74k | 183 (18.1) | 174 (17.2) | 57 (18.6) | 105 (15.9) | |
| \$75-99k | 147 (14.6) | 141 (14.0) | 32 (10.5) | 105 (16.0) | |
| \$100-124k | 108 (10.7) | 103 (10.3) | 33 (10.8) | 67 (10.2) | |
| \$125-149k | 73 (7.2) | 66 (6.5) | 112 (3.8) | 52 (7.9) | |
| \$150-174k | 90 (8.9) | 82 (8.1) | 25 (8.1) | 53 (8.0) | |
| \$175k+ | 140 (13.9) | 124 (12.3) | 32 (10.4) | 86 (13.1) | |
| **Education** |  |  |  |  | 0.1 |
| Less than high school | 75 (7.4) | 98 (9.7) | 39 (12.7) | 52 (7.9) |  |
| High school | 278 (27.5) | 295 (29.2) | 83 (27.4) | 186 (28.4) |  |
| Some college | 273 (27.0) | 281 (27.8) | 85 (28.1) | 189 (28.7) |  |
| Bachelor's degree or higher | 384 (38.0) | 336 (33.3) | 97 (31.9) | 230 (35.0) |  |
| **Children in home (<13 years)** |  |  |  |  | 1.0 |
| Yes | 194 (19.2) | 223 (22.0) | 67 (22.0) | 145 (22.0) |  |
| No | 816 (80.8) | 787 (78.0) | 238 (78.0) | 512 (78.0) |  |
| **Living alone** |  |  |  |  | 0.05 |
| Yes | 190 (18.8) | 184 (18.2) | 66 (21.5) | 107 (16.3) |  |
| No | 820 (81.2) | 826 (81.8) | 239 (78.5) | 550 (83.7) |  |
| **Household size** |  |  |  |  | 0.4 |
| Mean (SD) | 2.65 (1.50) | 2.77 (1.60) | 2.72 (1.66) | 2.81 (1.58) |  |
| **Loneliness scale<sup>2</sup>** |  |  |  |  | <0.0001 |
| Mean (SD) | 4.31 (1.69) | 4.37 (1.74) | 5.48 (2.08) | 3.90 (1.33) |  |

<sup>1</sup>Major depressive symptoms measured with the CES-D-10 scale. Possible scores range 0-30. The frequency of missing data for this variable was n=44 (4.4%).
<sup>2</sup>Loneliness measured with the UCLA 3-Item Loneliness scale. Possible scores ranged 3-9. The frequency of missing observations for this variable was n=19 (1.9%)

The weighted prevalence of major depressive symptoms in the last week was 31.7% (95% CI: 28.9, 34.8%). People reporting major depressive symptoms were more likely to be women, age 20–29, unmarried, in the lowest income bracket, and living alone. Loneliness scale scores were also significantly higher among those with major depressive symptoms [mean (SD): 5.5 (2.1)], than among those without [mean (SD) 3.9 (1.3)]. The weighted prevalence of loneliness was 54.0% (95% CI: 51.0, 57.2%) using our first definition, and 23.8% (95% CI: 21.3, 26.6%) using our second definition.

In general, frequent in-person social connections were associated with lower depression and loneliness prevalence, but frequent remote social connections were not (Table 2, Figure 1). Compared to those who reported ‘not at all,’ people who hugged or kissed a family member almost every day in last month were 26% less likely to report major depressive symptoms [aPR (95% CI): 0.74 (0.56, 0.97)], and 28% less likely to report loneliness [aPR (95% CI): 0.72 (0.51, 1.02)]. Though not statistically significant, we observed similar trends for the exposure of in-person visits with non-household members [aPRdepression (95% CI): 0.79 (0.48, 1.31) and aPRloneliness (95% CI): 0.86 (0.51, 1.02)]. This trend was not observed for remote social connections (video chats and sending/receiving letters in the mail). Relative to ‘not at all’ responses, high frequency of video chats were not associated with lower depression or loneliness prevalence [aPR (95% CI): 1.02 (0.78, 1.35) and aPR (95% CI): 1.12 (0.79, 1.59), respectively], whereas those with high frequency of sending/receiving letters tended towards higher depression and loneliness [aPR (95% CI): 1.57 (0.95, 2.59) and aPR (95% CI): 2.49 (1.33, 4.65), respectively].

**Table 2.**
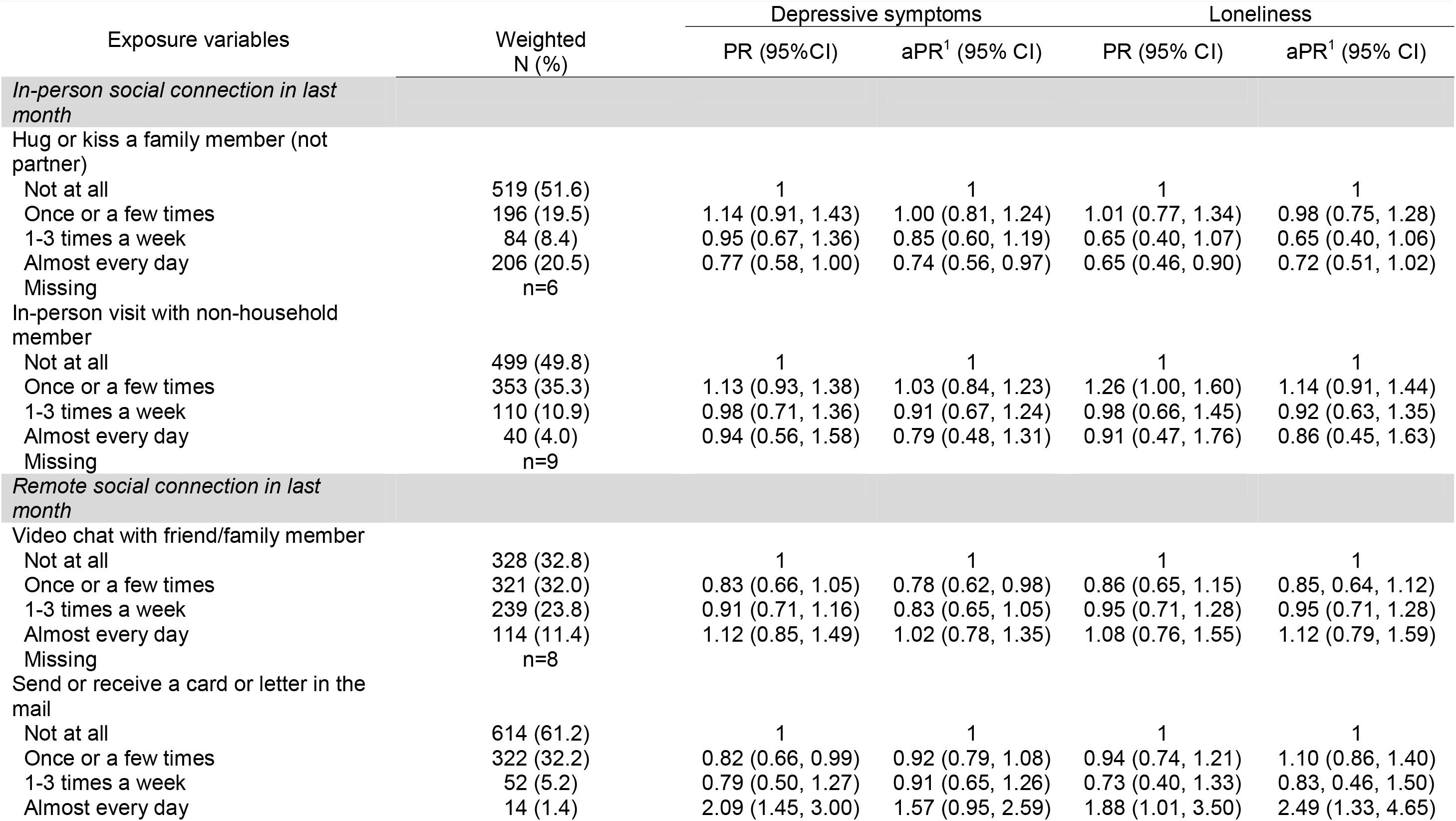
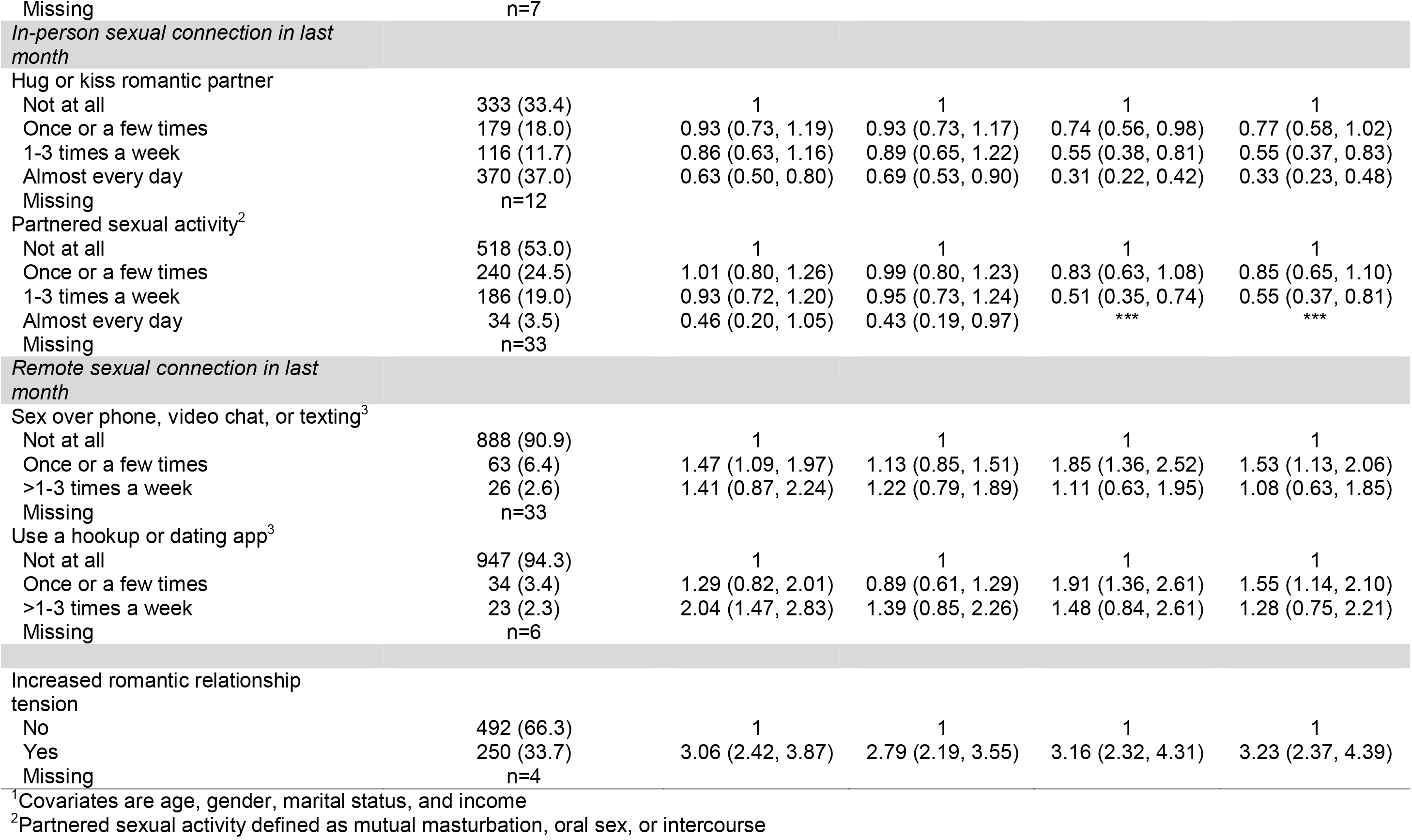
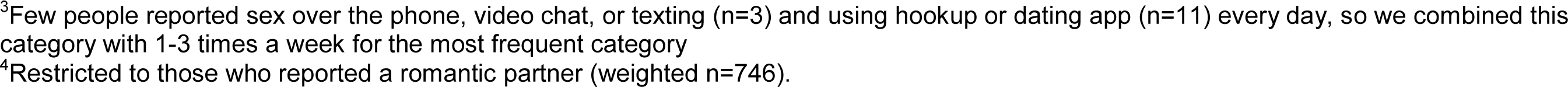
Unadjusted and adjusted log-binomial regression for the associations between frequency of social and sexual connections and outcomes of depression and loneliness

| Exposure variables | Weighted<br>N (%) | Depressive symptoms |  | Loneliness |  |
| --- | --- | --- | --- | --- | --- |
|  |  | PR (95%CI) | aPR<sup>1</sup> (95% CI) | PR (95% CI) | aPR<sup>1</sup> (95% CI) |
| *In-person social connection in last month* |  |  |  |  |  |
| Hug or kiss a family member (not partner) |  |  |  |  |  |
| Not at all | 519 (51.6) | 1 | 1 | 1 | 1 |
| Once or a few times | 196 (19.5) | 1.14 (0.91, 1.43) | 1.00 (0.81, 1.24) | 1.01 (0.77, 1.34) | 0.98 (0.75, 1.28) |
| 1-3 times a week | 84 (8.4) | 0.95 (0.67, 1.36) | 0.85 (0.60, 1.19) | 0.65 (0.40, 1.07) | 0.65 (0.40, 1.06) |
| Almost every day | 206 (20.5) | 0.77 (0.58, 1.00) | 0.74 (0.56, 0.97) | 0.65 (0.46, 0.90) | 0.72 (0.51, 1.02) |
| Missing | n=6 |  |  |  |  |
| In-person visit with non-household member |  |  |  |  |  |
| Not at all | 499 (49.8) | 1 | 1 | 1 | 1 |
| Once or a few times | 353 (35.3) | 1.13 (0.93, 1.38) | 1.03 (0.84, 1.23) | 1.26 (1.00, 1.60) | 1.14 (0.91, 1.44) |
| 1-3 times a week | 110 (10.9) | 0.98 (0.71, 1.36) | 0.91 (0.67, 1.24) | 0.98 (0.66, 1.45) | 0.92 (0.63, 1.35) |
| Almost every day | 40 (4.0) | 0.94 (0.56, 1.58) | 0.79 (0.48, 1.31) | 0.91 (0.47, 1.76) | 0.86 (0.45, 1.63) |
| Missing | n=9 |  |  |  |  |
| *Remote social connection in last month* |  |  |  |  |  |
| Video chat with friend/family member |  |  |  |  |  |
| Not at all | 328 (32.8) | 1 | 1 | 1 | 1 |
| Once or a few times | 321 (32.0) | 0.83 (0.66, 1.05) | 0.78 (0.62, 0.98) | 0.86 (0.65, 1.15) | 0.85, 0.64, 1.12) |
| 1-3 times a week | 239 (23.8) | 0.91 (0.71, 1.16) | 0.83 (0.65, 1.05) | 0.95 (0.71, 1.28) | 0.95 (0.71, 1.28) |
| Almost every day | 114 (11.4) | 1.12 (0.85, 1.49) | 1.02 (0.78, 1.35) | 1.08 (0.76, 1.55) | 1.12 (0.79, 1.59) |
| Missing | n=8 |  |  |  |  |
| Send or receive a card or letter in the mail |  |  |  |  |  |
| Not at all | 614 (61.2) | 1 | 1 | 1 | 1 |
| Once or a few times | 322 (32.2) | 0.82 (0.66, 0.99) | 0.92 (0.79, 1.08) | 0.94 (0.74, 1.21) | 1.10 (0.86, 1.40) |
| 1-3 times a week | 52 (5.2) | 0.79 (0.50, 1.27) | 0.91 (0.65, 1.26) | 0.73 (0.40, 1.33) | 0.83, 0.46, 1.50) |
| Almost every day | 14 (1.4) | 2.09 (1.45, 3.00) | 1.57 (0.95, 2.59) | 1.88 (1.01, 3.50) | 2.49 (1.33, 4.65) |
| Missing | n=7 |  |  |  |  |
| *In-person sexual connection in last month* |  |  |  |  |  |
| Hug or kiss romantic partner |  |  |  |  |  |
| Not at all | 333 (33.4) | 1 | 1 | 1 | 1 |
| Once or a few times | 179 (18.0) | 0.93 (0.73, 1.19) | 0.93 (0.73, 1.17) | 0.74 (0.56, 0.98) | 0.77 (0.58, 1.02) |
| 1-3 times a week | 116 (11.7) | 0.86 (0.63, 1.16) | 0.89 (0.65, 1.22) | 0.55 (0.38, 0.81) | 0.55 (0.37, 0.83) |
| Almost every day | 370 (37.0) | 0.63 (0.50, 0.80) | 0.69 (0.53, 0.90) | 0.31 (0.22, 0.42) | 0.33 (0.23, 0.48) |
| Missing | n=12 |  |  |  |  |
| Partnered sexual activity<sup>2</sup> |  |  |  |  |  |
| Not at all | 518 (53.0) | 1 | 1 | 1 | 1 |
| Once or a few times | 240 (24.5) | 1.01 (0.80, 1.26) | 0.99 (0.80, 1.23) | 0.83 (0.63, 1.08) | 0.85 (0.65, 1.10) |
| 1-3 times a week | 186 (19.0) | 0.93 (0.72, 1.20) | 0.95 (0.73, 1.24) | 0.51 (0.35, 0.74) | 0.55 (0.37, 0.81) |
| Almost every day | 34 (3.5) | 0.46 (0.20, 1.05) | 0.43 (0.19, 0.97) | *** | *** |
| Missing | n=33 |  |  |  |  |
| *Remote sexual connection in last month* |  |  |  |  |  |
| Sex over phone, video chat, or texting<sup>3</sup> |  |  |  |  |  |
| Not at all | 888 (90.9) | 1 | 1 | 1 | 1 |
| Once or a few times | 63 (6.4) | 1.47 (1.09, 1.97) | 1.13 (0.85, 1.51) | 1.85 (1.36, 2.52) | 1.53 (1.13, 2.06) |
| >1-3 times a week | 26 (2.6) | 1.41 (0.87, 2.24) | 1.22 (0.79, 1.89) | 1.11 (0.63, 1.95) | 1.08 (0.63, 1.85) |
| Missing | n=33 |  |  |  |  |
| Use a hookup or dating app<sup>3</sup> |  |  |  |  |  |
| Not at all | 947 (94.3) | 1 | 1 | 1 | 1 |
| Once or a few times | 34 (3.4) | 1.29 (0.82, 2.01) | 0.89 (0.61, 1.29) | 1.91 (1.36, 2.61) | 1.55 (1.14, 2.10) |
| >1-3 times a week | 23 (2.3) | 2.04 (1.47, 2.83) | 1.39 (0.85, 2.26) | 1.48 (0.84, 2.61) | 1.28 (0.75, 2.21) |
| Missing | n=6 |  |  |  |  |
| *Increased romantic relationship tension* |  |  |  |  |  |
| No | 492 (66.3) | 1 | 1 | 1 | 1 |
| Yes | 250 (33.7) | 3.06 (2.42, 3.87) | 2.79 (2.19, 3.55) | 3.16 (2.32, 4.31) | 3.23 (2.37, 4.39) |
| Missing | n=4 |  |  |  |  |

<sup>1</sup>Covariates are age, gender, marital status, and income
<sup>2</sup>Partnered sexual activity defined as mutual masturbation, oral sex, or intercourse
<sup>3</sup>Few people reported sex over the phone, video chat, or texting (n=3) and using hookup or dating app (n=11) every day, so we combined this category with 1-3 times a week for the most frequent category
<sup>4</sup>Restricted to those who reported a romantic partner (weighted n=746).

**Figure 1.**
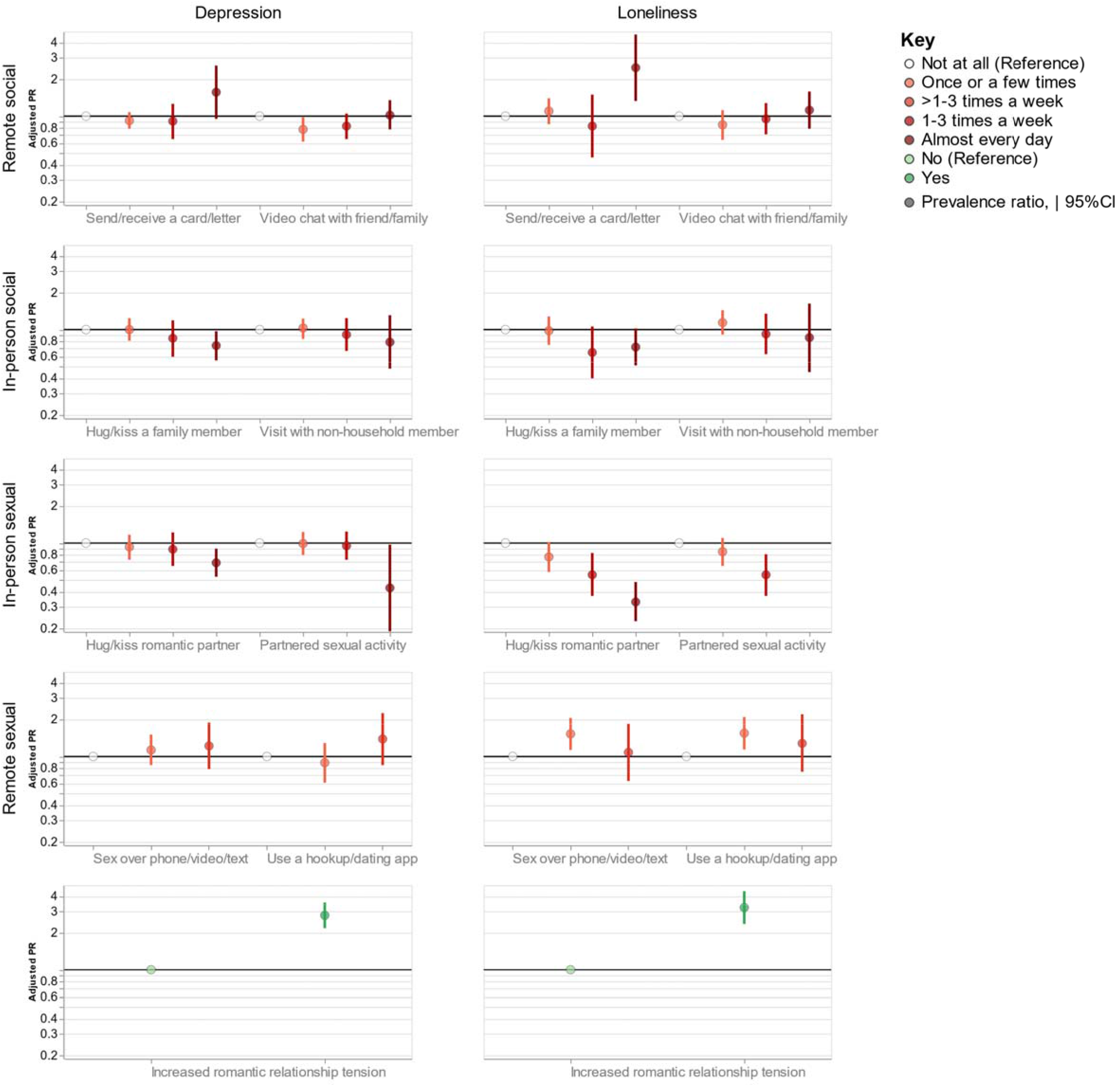
Associations between frequencies of social and sexual connections and outcomes of depressive e symptoms(top panel) and loneliness (bottom panel) in 1010 American adults, April 10–20, 2020.

Similarly, frequent in-person sexual connections were associated with lower prevalence of depression and loneliness in ways that remote sexual connections were not (Table 2, Figure 1). For the hug/kiss and partnered sex exposures, a dose response relationship was observed where, relative to ‘not at all’ responses, increasing frequencies were associated with decreased depression and loneliness prevalence. Those who endorsed the most frequent hug/kiss exposure were 31% less likely to report depressive symptoms and 67% less likely to report loneliness [aPR (95% CI): 0.69 (0.53, 0.90) and aPR (95% CI): 0.33 (0.23, 0.48), respectively]. Those who endorsed the most frequent partnered sex exposure were 57% less likely to report depressive symptoms [aPR (95% CI): 0.43 (0.19, 0.97)]. No participant with partnered sex at this high frequency had loneliness scores above 6. Relatively few participants endorsed frequent remote sex or dating app usage, but those who did tended to have slightly higher rates of depression and loneliness. However, these estimates were measured imprecisely with wide confidence limits spanning the null. Participants who endorsed experiencing increased tension with their romantic partner due to COVID-19 spread and restrictions were more likely to report major depressive symptoms [aPR (95% CI): 2.79 (2.19, 3.55)] and loneliness [aPR (95% CI): 3.23 (2.37, 4.39)].

The patterns for the associations between social and sexual connections and mental health outcomes generally did not differ depending on if the participant lived with a romantic partner (Table 3). One possible exception to this trend emerged for the remote sex over phone, video chat, or texting outcome, though the low numbers of participants reporting this exposure frequently limited the precision of our estimates. For participants who lived with a partner, more frequent remote sex was associated with lower depression and loneliness [aPR (95% CI): 0.89 (0.38, 2.07) and aPR (95% CI): 0.34 (0.04, 2.70), respectively], while the opposite was true for participants who did not live with a partner [aPR (95% CI): 1.66 (0.97, 2.84) and aPR (95% CI): 1.69 (0.97, 2.84), respectively, Wald p-values for interaction both=0.2].

**Table 3.** Adjusted log-binomial regression for the associations between frequency of social and sexual connections and outcomes of depression and loneliness, stratified by sexual relationship type (living with partner vs not living with partner)

| Exposure variables | Depressive symptoms |  |  | Loneliness |  |  |
| --- | --- | --- | --- | --- | --- | --- |
|  | Live with a partner<br>aPR<sup>1</sup> (95%CI) | Do not live with a partner<br>aPR<sup>1</sup> (95% CI) | p for interaction | Live with a partner<br>aPR<sup>1</sup> (95%CI) | Do not live with a partner<br>aPR<sup>1</sup> (95% CI) | p for interaction |
| *In-person social connection in last month* |  |  |  |  |  |  |
| Hug or kiss a family member (not partner) |  |  |  |  |  |  |
| <1-3 times a week | 1 | 1 |  | 1 | 1 |  |
| 1-3 times a week or more | 0.68 (0.50, 0.91) | 0.91 (0.66, 1.25) | 0.2 | 0.63 (0.41, 0.96) | 0.73 (0.50, 1.07) | 0.6 |
| In-person visit with non-household member |  |  |  |  |  |  |
| <1-3 times a week | 1 | 1 |  | 1 | 1 |  |
| 1-3 times a week or more | 0.94 (0.65, 1.37) | 0.85 (0.59, 1.24) | 0.7 | 0.91 (0.53, 1.55) | 0.77 (0.51, 1.18) | 0.6 |
| *Remote social connection in last month* |  |  |  |  |  |  |
| Video chat with friend/family member |  |  |  |  |  |  |
| <1-3 times a week | 1 | 1 |  | 1 | 1 |  |
| 1-3 times a week or more | 1.05 (0.81, 1.36) | 0.93 (0.72, 1.21) | 0.5 | 0.94 (0.65, 1.36) | 1.20 (0.91, 1.58) | 0.3 |
| Send or receive a card or letter in the mail |  |  |  |  |  |  |
| <1-3 times a week | 1 | 1 |  | 1 | 1 |  |
| 1-3 times a week or more | 1.23 (0.79, 1.91) | 1.47 (0.92, 2.34) | 0.6 | 1.39 (0.78, 2.48) | 1.00 (0.48, 2.07) | 0.5 |
| *In-person sexual connection in last month* |  |  |  |  |  |  |
| Hug or kiss romantic partner |  |  |  |  |  |  |
| <1-3 times a week | 1 | 1 |  | 1 | 1 |  |
| 1-3 times a week or more | 0.68 (0.52, 0.89) | 0.94 (0.64, 1.38) | 0.2 | 0.44 (0.31, 0.64) | 0.56 (0.32, 0.97) | 0.5 |
| Partnered sexual activity** |  |  |  |  |  |  |
| <1-3 times a week | 1 | 1 |  | 1 | 1 |  |
| 1-3 times a week or more | 0.81 (0.61, 1.08) | 1.12 (0.69, 1.82) | 0.3 | 0.48 (0.30, 0.76) | 0.71 (0.35, 1.43) | 0.4 |
| *Remote sexual connection in last month* |  |  |  |  |  |  |
| Sex over phone, video chat, or texting*** |  |  |  |  |  |  |
| <1-3 times a week | 1 | 1 |  | 1 | 1 |  |
| 1-3 times a week or more | 0.89 (0.38, 2.07) | 1.66 (0.97, 2.84) | 0.2 | 0.34 (0.04, 2.70) | 1.45 (0.85, 2.48) | 0.2 |
| Use a hookup or dating app*** |  |  |  |  |  |  |
| <1-3 times a week | 1 | 1 |  | 1 | 1 |  |
| 1-3 times a week or more | 1.61 (0.88, 2.95) | 1.67 (1.13, 2.48) | 0.9 | 1.79 (0.71, 4.49) | 2.28 (1.62, 3.22) | 0.6 |
| Increased romantic relationship tension<sup>2</sup> |  |  |  |  |  |  |
| No | 1 | 1 |  | 1 | 1 |  |
| Yes | 3.16 (2.39, 4.17) | 2.00 (1.30, 3.08) | 0.08 | 3.26 (2.23, 4.74) | 3.27 (1.95, 5.47) | 1.0 |

<sup>1</sup>Covariates are age, gender, and income. Marital status excluded because of overlapping definition with stratifying variable.
<sup>2</sup>Restricted to those who reported a romantic partner (weighted n=746).

## DISCUSSION

In a nationally representative probability survey of American adults taking place during the early phases of the COVID-19 response (April 2020), we found high levels of significant depressive symptoms and loneliness. Frequent in-person social and sexual contacts during the time of COVID-19 restrictions in the last month were generally associated with lower prevalence of depression and loneliness, while remote contacts were not similarly protective. We also found that relationship tension due to COVID-19 spread and restrictions was strongly predictive of depression and loneliness, and that these associations were significantly stronger among people who lived with their romantic partners. Our findings suggest that the COVID-19 spread and response has had a tremendous mental health impact on Americans.

We found that nearly one-third of Americans reported depressive symptoms in April 2020, notably much higher than previous estimates among American adults. From 2013–2016, the prevalence of major depression in a given 2 week period in US adults age 20+ years was 8%, nearly 4 times lower than our estimate.[30] Similarly, the mean loneliness score we found (4.4) was higher than prior estimates in three western European countries (3.5–3.7),[31] and the prevalence of loneliness we found in our general US population (Definition 1: 54%), is a similar magnitude as previously observed in older and elderly Americans (43–56%).[28, 29] Patterns of higher depression prevalence among women, young adults, and people with lower incomes have been observed in previous studies.[14, 30] We also observed these gender, age, and socioeconomic patterns in our study, suggesting that while the magnitude of depression may be expanding during the time of COVID-19 spread and restrictions, the disproportionate burden continues to be felt in these populations.

The observed relationships between social and sexual connections and the outcomes of depression and loneliness are largely consistent with our understanding of the importance of human connection for mental health and well-being. Close touching in family interactions and among relationship partners has been associated with decreased heart rate, higher levels of oxytocin, and lower levels of cortisol;[32, 33] which, in turn could provide important mental health protections. These kinds of connections cannot easily be recreated with remote technology where direct touch is not possible, though some researchers have explored this through “huggable” and other similar communication devices.[34] We also know that common technical difficulties with video calls can cause misattributed negative feelings towards people on the call,[35] perhaps contributing to the null effects we observed for remote connections and mental health outcomes. As the COVID-19 spread and response have dramatically limited access to many previously routine and familiar options for human connection, our findings are consistent with an explanation that the decreased frequency of social and sexual connections is contributing to poor mental health outcomes. In addition to limiting people’s social interactions, restrictions have also likely resulted in many people missing counselling or therapy appointments or other activities (e.g., exercise, massage, support group meetings, etc.) that may have been supportive of their mental health.

There are several other plausible explanations for the observed relationship between social and sexual connections and mental health outcomes. First, people who are struggling with poor mental health and feelings of isolation during COVID-19 restrictions may be reaching out more frequently for remote connections. As this was a cross-sectional study, our ability to infer temporality is limited. Future studies with a longitudinal design would be better able to disentangle the temporal relationships between social/sexual connections and mental health outcomes. Second, we used self-reported data for our key exposure and outcome assessments, with the potential for biased results. However, web-based surveys have shown to be effective modes to elicit sensitive sexual behaviors from study participants, alleviating some of the bias concern.[36] Similarly, our depression and loneliness scales are widely used in web-based surveys, are validated, and have good psychometric properties.[24, 27] However, their use could be complemented with clinical assessments in future studies. Relatedly, it is not yet known how these scales perform during times of social restrictions when daily lives have shifted so dramatically. It is possible that the wording of certain items (e.g. ‘How often do you feel isolated from others?’) may lose their discriminatory ability in times where isolation is essentially mandated. For this reason, we used a more conservative categorical definition of loneliness (scale score of 6 or greater) for our analyses than has been commonly used in prior studies (scale score of 4 or greater). We also did not explore whether participants had run out of prescription medication supportive of their mental health or had missed counselling or therapy visits.

A key strength of this study is its external validity. Our findings are broadly representative of the US adult population during the early stages of the US COVID-19 response. However, these results may not hold during later phases of the COVID-19 response, or for countries outside of the US. Surveys should be conducted at frequent intervals and in other countries affected by COVID-19 to update our understanding of the changing mental health landscape across time and space.

### Conclusions

Depressive symptoms and loneliness are being experienced at very high levels in Americans during the COVID-19 spread and response. At this time, social restrictions are a critical tool to mitigate the spread of COVID-19 and its associated morbidity and mortality. Our findings should not be interpreted in support of prematurely lifting these necessary restrictions. Rather, public health and mental health professionals should be aware of the increasing mental health needs that are likely co-occurring with the COVID-19 response, and funding should be allocated to expand mental health services to those most likely to be experiencing poor mental health. Women, young adults, and low-income Americans seem to be particularly at-risk for these outcomes. We also found that Americans who could maintain frequent in-person social and sexual connections appeared to have better mental health outcomes, though remote connections did not appear to confer the same benefits. Given that extra-household connections are not advised at this time and possibly will be contraindicated intermittently in the future, a mental health priority should be to identify innovative and effective ways of supporting interpersonal connections from a distance.

## Data Availability

Data and code underlying the findings presented in this paper are available from the first author upon request.

## DECLARATIONS

### Ethics approval

Ethical approval for the study protocol was provided by the Indiana University Human Subjects Office (#2004194314).

### Authors’ contributions

M. Rosenberg and D. Herbenick conceived the study. D. Herbenick, M. Rosenberg, M. Luetke, and D. Hensel designed the survey. M. Rosenberg conducted the analysis and wrote the first draft of the manuscript with key drafting contributions from ML and scientific contributions from D. Hensel, T. Fu, and D. Herbenick. S. Kianersi created the data visualization. M. Rosenberg, M. Luetke, D. Hensel, T. Fu, S. Kianersi, and D. Herbenick contributed to the interpretation of the findings, critical review of the manuscript, and approval of the final manuscript as submitted.

